# Risk-Based Cardiovascular-Kidney-Metabolic Staging Identifies a Population With Mortality Comparable to Established Cardiovascular Disease: Disability-Adjusted Life Years Across AHA Stages, 2005 to 2023

**DOI:** 10.64898/2026.11.05.26367395

**Authors:** Xiaolei Wang, Yu Qiao, Hongfeng Sun, Ying Wang, Hui Zhou, Taojing Zhang

## Abstract

**Background:** CKM syndrome was defined in 2023 as a five-stage continuum, with stage 3 assigned on predicted rather than established cardiovascular disease. Whether that stage identifies a population with mortality resembling clinical disease is unknown.

**Methods:** We pooled nine National Health and Nutrition Examination Survey cycles (2005-2023) and assigned CKM stages 0-4 from measured risk factors, kidney function, urine albumin, self-reported cardiovascular disease, and 10-year PREVENT risk. Years lived with disability used Global Burden of Disease weights; years of life lost combined national deaths, linked mortality files, and period life tables.

**Results:** In 2021-2023, 13.8%, 21.3%, 49.1%, 6.5%, and 9.3% of adults were in stages 0 to 4. Crude mortality was 6,709 per 100,000 person-years in stage 3 and 6,375 in stage 4. Adjusted hazard ratios increased from 1.39 in stage 2 to 3.22 in stage 4; stage 1 showed no significant excess (HR, 0.99). CKM accounted for 20.0 million DALYs annually, with 76% of attributable years of life lost in stages 3-4. Lipid-lowering therapy was absent in 53.7% of stage 3 and 33.6% of stage 4 adults, a gap that did not narrow over 16 years.

**Conclusions:** Risk-based stage 3 had a crude mortality rate similar to that of established cardiovascular disease, although its adjusted hazard ratio remained lower. The population nonetheless shifted toward stage 1, and stage 3 continued to receive less intensive lipid-lowering treatment than stage 4.

**Clinical Perspective:** *What Is New?:*

- First quantification of disability-adjusted life years across all five AHA CKM stages. Risk-based stage 3 had a crude mortality rate similar to that of stage 4 (6,709 versus 6,375 per 100,000 person-years), although its adjusted hazard ratio remained lower (2.21 versus 3.22).
- Stage 1 was not associated with a statistically significant excess in adjusted mortality (HR 0.99; 95% CI, 0.78-1.25); excess risk became significant at stage 2. Stage 1 nonetheless expanded by 3.1% per year between 2006 and 2016.
- Stages 3 and 4 generate 76% of attributable years of life lost, yet adults in stage 3 were 20 percentage points more likely to be untreated for lipid risk, a gap that has not narrowed since 2005.

*What Are the Clinical Implications?:*

- Mortality-focused case finding may yield the greatest short-term benefit in stages 3-4. Management of stages 1 and 2 should also consider the prevention of progression, not solely short-term mortality benefit.
- The absence of a statistically significant excess in adjusted mortality in stage 1 may help clinicians communicate population-level risk findings and support engagement in primordial prevention.
- Because burden metrics emphasize late-stage outcomes, CKM programs that rely on DALY-based prioritization may underinvest in earlier stages where the population is accumulating.

## Introduction

In 2023, the American Heart Association proposed cardiovascular-kidney-metabolic (CKM) syndrome as a single construct linking adiposity, dysglycemia, hypertension, dyslipidemia, chronic kidney disease, and cardiovascular disease along a five-stage continuum from stage 0 (no risk factors) to stage 4 (clinical cardiovascular disease).1 The framework has been rapidly adopted in epidemiologic research, and national estimates of stage distribution are now available.2

Stage 3 is the least tested part of the framework. Unlike the other stages, it is assigned on predicted ten-year risk rather than on measured disease, and it combines adults with a PREVENT score of 20% or more and adults with a high-risk kidney profile. Whether this group has mortality similar to that of adults with established cardiovascular disease has not been tested against national death data. If it does, the stage identifies a group that warrants intensive prevention; if it does not, the label has limited prognostic value.

National estimates describe how many people occupy each stage, but not how much health they lose. A syndrome whose burden sits in advanced stages argues for case finding and secondary prevention; one whose burden sits in early stages argues for primordial prevention. Prevalence alone cannot adjudicate between the two, because a common early stage and a rare late stage can carry identical prevalence-weighted risk.

Disability-adjusted life years (DALYs) resolve this by combining years lived with disability (YLD) and years of life lost (YLL), and by permitting decomposition across stages. DALYs are the standard metric of the Global Burden of Disease (GBD) study,^3^ but to our knowledge they have never been estimated by CKM stage. Existing CKM work has examined single biomarkers across stages,^4,5^ albuminuria prevalence,^6^ and mortality,^7^ but not health loss.

We therefore estimated years lived with disability, years of life lost, and DALYs for each CKM stage among US adults from 2005 to 2023, and tested the mortality gradient that the framework implies. We had three questions. First, does a PREVENT-defined stage 3 carry mortality comparable to stage 4? Second, how is health loss distributed across stages, and does it sit where the deaths are? Third, how has the stage distribution moved over two decades, and does that movement point upstream or downstream of where the burden sits?

## Methods

### Data sources and study population

We used nine National Health and Nutrition Examination Survey (NHANES) cycles: 2005-2006 through 2017-2018, the 2017-March 2020 pre-pandemic cycle, and 2021-2023. NHANES is a stratified, multistage probability sample of the noninstitutionalized US population.8 We restricted the prevalence and YLD analyses to adults aged 20 years or older who attended the mobile examination center and had a positive examination weight (Figure S1); variable definitions and source files for each cycle are listed in Table S2. The 2017-2018 cycle was retained for internal consistency checks but excluded from trend analyses because it overlaps the 2017-March 2020 cycle.

Because triglycerides are measured only in the fasting subsample, the prevalence and YLD analyses were restricted to fasting participants. The corresponding fasting subsample weight (WTSAF2YR; WTSAFPRP for 2017-March 2020) was applied to these weighted analyses. The analytic sample comprised 3,057 adults in 2021-2023 and 23,951 across all cycles. Mortality analyses used a separate pooled linked-mortality cohort of 27,012 participants, as described below.

### CKM staging

Stages were assigned hierarchically, with the highest applicable stage taking precedence (Table S1). Stage 4 required self-reported congestive heart failure, coronary heart disease, angina, myocardial infarction, or stroke. Stage 3 required the absence of clinical cardiovascular disease plus either a 10-year PREVENT total cardiovascular risk of 20% or greater or high-risk chronic kidney disease (estimated glomerular filtration rate [eGFR] <45 mL/min/1.73 m² or urine albumin-to-creatinine ratio [UACR] ≥300 mg/g). Stage 2 required any of the following: hypertension, diabetes, chronic kidney disease, hypertriglyceridemia, metabolic syndrome, or low-density lipoprotein cholesterol ≥160 mg/dL. Stage 1 required overweight, obesity, abdominal obesity, or prediabetes. Stage 0 required none of the above.

Hypertension was defined by measured systolic blood pressure >=130 mm Hg or diastolic >=80 mm Hg, or current antihypertensive medication; self-report of a prior diagnosis was deliberately excluded so that treated and controlled individuals were classified by current status. Diabetes was defined by self-report, glucose-lowering medication, or hemoglobin A1c >=6.5%. Prediabetes was defined as hemoglobin A1c 5.7%-6.4% without diabetes. Kidney function was estimated with the 2021 CKD-EPI creatinine equation.^9^ Stage 0 was assigned only when all five core inputs (body mass index, blood pressure, hemoglobin A1c, eGFR, and UACR) were observed, so that missing data could not create spurious health.

Ten-year cardiovascular risk was computed with the PREVENT base equations,^10^ implemented from the published coefficients and validated against the reference implementation. PREVENT was calculable for 75% of adults; participants younger than 30 years or with incomplete inputs could not meet the stage 3 risk criterion.

### Disability weights and YLD

YLD were calculated as the product of stage-specific prevalence, disability weight, and population size. Disability weights were assigned at the level of GBD health states rather than conditions. Diabetes was assigned a weight of 0.049. For ischemic heart disease and chronic ischemic stroke, we applied GBD severity distributions11 to obtain severity-weighted averages of 0.0731 and 0.0845, respectively. Heart failure used the GBD 2023 severity distribution (0.0923).12 Chronic kidney disease weights were stratified by anemia severity using hemoglobin and the GBD CKD severity table: for stage 3, weights were 0 without anemia and 0.004, 0.052, and 0.149 with mild, moderate, and severe anemia, respectively; for stage 4, weights ranged from 0.104 to 0.237; and for stage 5 (dialysis), the weight was 0.569 (Table S5).13 Coexisting conditions were combined multiplicatively, DW = 1 - product(1 - DWi), as in the GBD approach to comorbid sequelae. Stages 0 and 1 were assigned zero disability because adiposity, prediabetes, and hypertension are treated by GBD as risk factors without an associated sequela.

### Mortality, YLL, and attributable burden

Vital status through 31 December 2019 was obtained from the public-use NHANES linked mortality files (LMF) for cycles 2005-2006 through 2017-2018, linked by participant sequence number.^14^ We pooled these cycles, together with the 1999-2000 and 2003-2004 cycles (2001-2002 was excluded because serum creatinine was not available in the public biochemistry file), to estimate the stage distribution of deaths within ten-year age bands and sex. Because the 1999-2000 and 2003-2004 cycles carry no survey weights, these shares were computed as unweighted counts of deaths within each age band and sex before being applied to national death counts. National death counts for 2021-2023 by year, sex, and ten-year age band were obtained from the Centers for Disease Control and Prevention WONDER database. Death counts for the three years were summed to give cumulative YLL over 2021-2023, and annual YLL was obtained by dividing that cumulative total by three; annual attributable YLL was then estimated by applying the PAF to the annualized total. Stage-specific deaths were obtained by applying the LMF-derived stage distribution of deaths to national counts; the 15-24 year band was halved to approximate ages 20-24. YLL were computed as deaths multiplied by remaining life expectancy from the 2024 US period life tables.^15^

Attributable burden was estimated with the population attributable fraction. Stage-specific hazard ratios (HR) were obtained from a Cox proportional hazards model on the pooled LMF cohort (27,012 participants; 4,706 deaths) with stage, age, sex, and race as covariates. The PAF was calculated as sum(pi(HRi-1)) / (1 + sum(pi(HRi-1))) using 2021-2023 stage prevalences, and attributable YLL were obtained by applying the PAF to total YLL, with the stage decomposition following each stage’s excess share. Cardiovascular deaths were identified from the underlying cause of death recode in the LMF (diseases of the heart or cerebrovascular disease), and cause-specific hazard ratios were estimated with deaths from other causes censored. The model used the same 27,012 participants as the all-cause model, among whom 1,530 cardiovascular deaths occurred. Rates were directly standardized to the 2020 US population.

### Attribution to upstream exposures

Smoking status and physical inactivity lie outside the CKM definition and were therefore treated as upstream exposures rather than components of the syndrome. Current smoking was defined by self-report. Physical inactivity was defined as reporting no leisure-time moderate or vigorous activity; the item set changed in 2007, and the two instruments were harmonized into a single binary indicator. Both exposures were available in every cycle from 2005-2006 onwards.

Hazard ratios were estimated in the same linked mortality cohort, with additional adjustment for the ratio of family income to poverty, educational attainment, and body mass index, so that the estimates would not merely reproduce socioeconomic position or adiposity. The population attributable fraction for the two exposures jointly was computed from their observed joint distribution in 2017-March 2020 and the adjusted hazard ratios for the four exposure combinations, using PAF = (sum p_s RR_s - 1) / sum p_s RR_s, which accounts for the overlap between exposures. Attributable DALYs were obtained by applying this fraction to total CKM DALYs.

### Statistical analysis

All prevalence estimates were survey-weighted using Taylor series linearization to account for strata and primary sampling units. Trends were modeled with log-linear segmented (joinpoint) regression, with up to two breakpoints selected by the Bayesian information criterion. Annual percentage change (APC) was reported for each segment, together with the average annual percentage change. Because NHANES changed from mercury sphygmomanometry to oscillometric devices in 2017, the primary trend analysis was restricted to 2006-2016, when the measurement protocol was constant; later cycles are shown but were not used for inference.

Uncertainty in the PAF and DALY estimates was quantified by Monte Carlo simulation (4,000 draws), combining the covariance of the Cox model with multinomial sampling of stage prevalence. Five prespecified sensitivity analyses were performed: (1) a PREVENT threshold of 30% rather than 20%; (2) omission of the low-density lipoprotein criterion; (3) relaxation of the stage 0 completeness rule; (4) disability weights set to the mild or severe end of the GBD severity range; and (5) alternative sources for the stage-specific distribution of deaths. Analyses were conducted using Python 3.13 (pandas, lifelines, statsmodels, and matplotlib). This study is reported in accordance with the Strengthening the Reporting of Observational Studies in Epidemiology (STROBE) guidelines for cross-sectional studies; the completed checklist is provided as supplementary material.

## Results

### Stage distribution

In 2021-2023, 13.8% (95% CI, 11.7-15.9) of US adults were stage 0, 21.3% (19.3-23.4) stage 1, 49.1% (46.6-51.6) stage 2, 6.5% (5.3-7.7) stage 3, and 9.3% (7.8-10.7) stage 4 (Table 1). Stage 2 was the largest category and was driven predominantly by hypertension (69.1%) and diabetes (16.7%). Stage 3 and stage 4 differed in composition: participants in stage 3 were older (mean 72.7 versus 66.0 years) and had worse kidney function (mean eGFR 70.0 versus 78.5 mL/min/1.73 m2) and more chronic kidney disease (55.5% versus 37.9%).

**Table 1.** Characteristics of US adults by CKM stage, NHANES 2021-2023. Values are survey-weighted means or percentages. PREVENT indicates 10-year predicted total cardiovascular risk.

| **Characteristic** | **Stage 0** | **Stage 1** | **Stage 2** | **Stage 3** | **Stage 4** |
|---|---|---|---|---|---|
| Participants, n | 320 | 557 | 1,514 | 282 | 384 |
| Age, mean, y | 36.4 | 40.4 | 49.7 | 72.7 | 66.0 |
| Female, % | 55.9 | 57.7 | 49.8 | 40.7 | 41.0 |
| Body mass index, mean | 22.0 | 30.3 | 31.4 | 30.2 | 31.0 |
| Systolic BP, mean, mm Hg | 109.8 | 110.2 | 124.1 | 135.7 | 126.1 |
| Haemoglobin A1c, mean, % | 5.2 | 5.3 | 5.9 | 6.3 | 6.3 |
| eGFR, mean, mL/min/1.73 m2 | 107.3 | 103.7 | 96.8 | 70.0 | 78.5 |
| Diabetes, % | 0.0 | 0.0 | 16.7 | 40.3 | 40.5 |
| Hypertension, % | 0.0 | 0.0 | 69.1 | 85.8 | 77.4 |
| Chronic kidney disease, % | 0.0 | 0.0 | 12.2 | 55.5 | 37.9 |
| Clinical CVD, % | 0.0 | 0.0 | 0.0 | 0.0 | 100.0 |
| PREVENT 10-year risk, mean, % | 2.4 | 3.2 | 7.4 | 25.2 | 17.9 |
| Mean disability weight | 0.000 | 0.000 | 0.009 | 0.034 | 0.126 |

### Disability and mortality burden

CKM accounted for 4,093,294 YLD and 15.9 million attributable YLL per year, or 20.0 million DALYs (8,537 per 100,000 adults crude; 7,881 age-standardized) (Table 2, Figure 2). YLL accounted for 80% of the total, indicating that CKM in the United States is principally a mortality burden, not a disability burden. Years of life lost per death fell from 29.8 in stage 0 to 10.1 in stage 3 and 13.1 in stage 4, the lowest value occurring in stage 3, whose members were the oldest.

**Figure 1.**
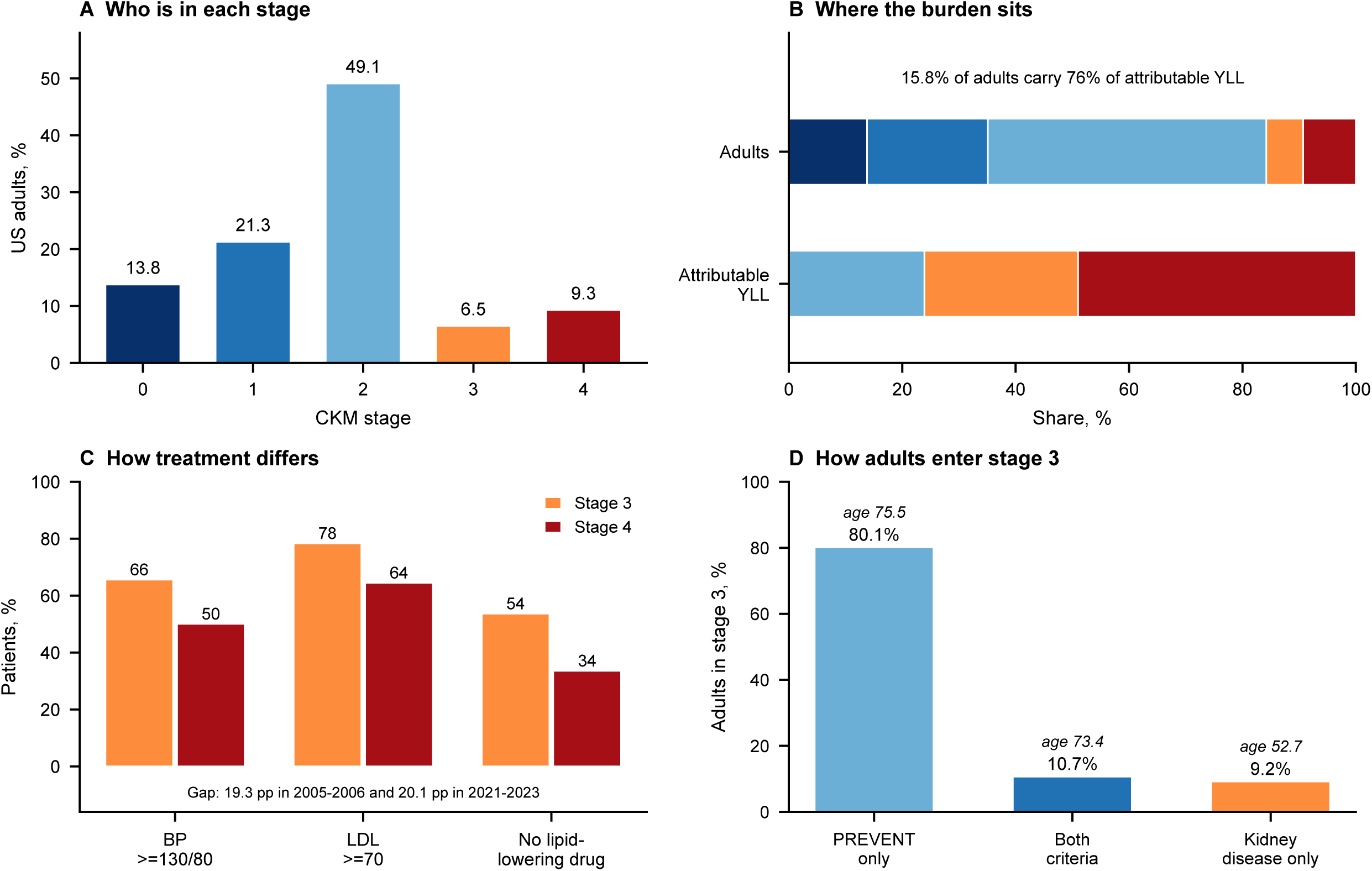
Survey-weighted prevalence of CKM stages among US adults, NHANES 2005-2023. Points are cycle midpoints. The shaded region marks cycles after the 2017 change from mercury sphygmomanometry to oscillometric blood pressure measurement; trends are inferred only from 2006-2016.

**Figure 2.**
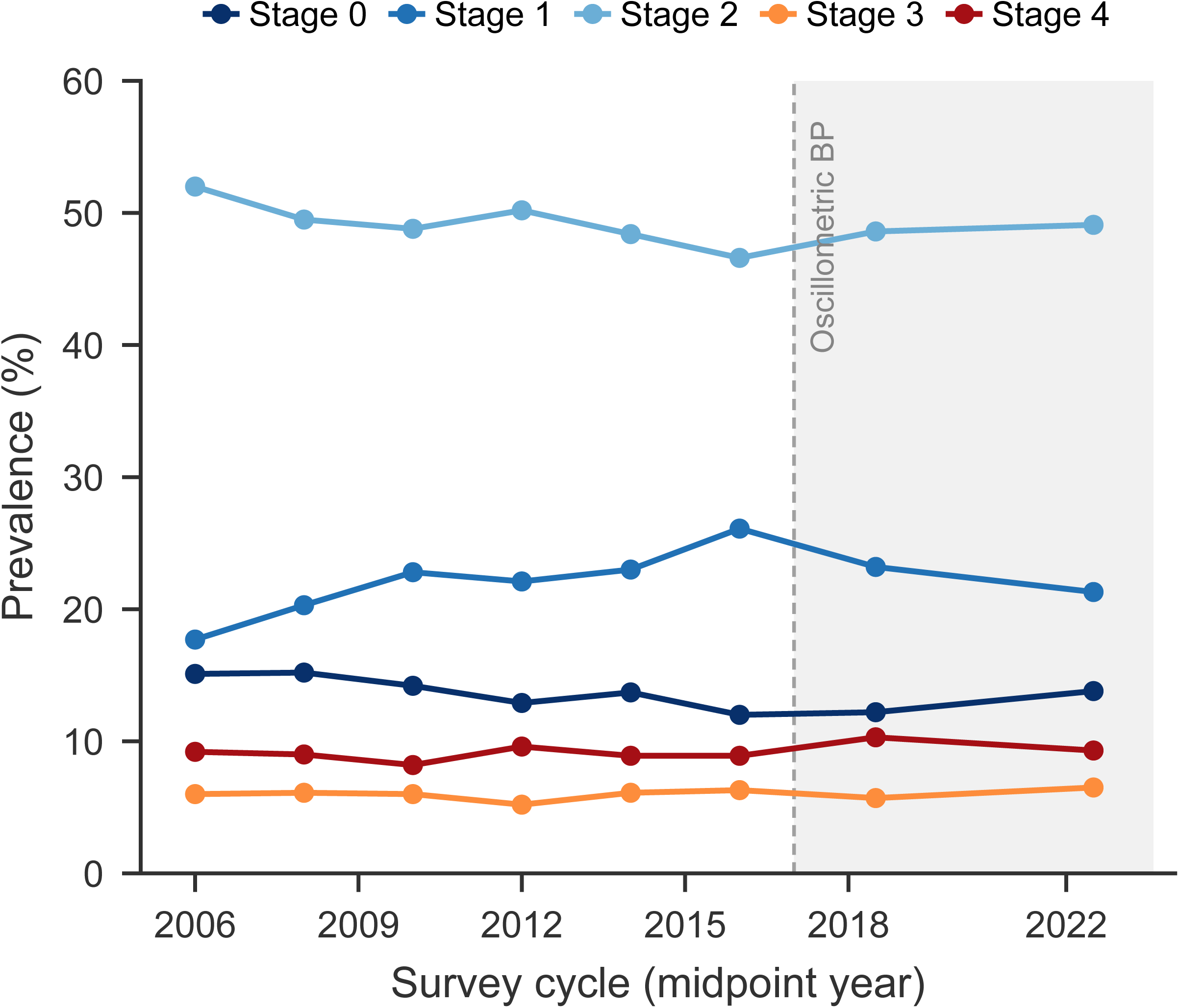
Annual DALYs by CKM stage, 2021-2023, decomposed into years lived with disability and attributable years of life lost. Stage 4 accounts for 52% of total DALYs.

**Table 2.** Annual YLD, attributable YLL, and DALYs by CKM stage, 2021-2023.

| **Stage** | **YLD** | **Attributable YLL** | **DALYs** | **DALYs per 100,000** | **% of DALYs** |
|---|---|---|---|---|---|
| 0 | 0 | 0 | 0 | 0 | 0.0 |
| 1 | 0 | 0 | 0 | 0 | 0.0 |
| 2 | 946,854 | 3,795,000 | 4,741,854 | 2,026 | 23.7 |
| 3 | 465,588 | 4,308,000 | 4,773,588 | 2,040 | 23.9 |
| 4 | 2,680,853 | 7,782,000 | 10,462,853 | 4,471 | 52.4 |
| **Total** | **4,093,294** | **15,885,000** | **19,978,295** | **8,537** | **100.0** |

Burden was concentrated in the advanced stages (Figure 2, Central Illustration). Stage 4 generated 65% of YLD and 49.0% of attributable YLL; stage 3 generated 11.4% and 27.1%; stage 2 generated 23.2% and 23.9%. Stages 3 and 4 together, 15.8% of adults, accounted for 76% of attributable YLL. The population attributable fraction for CKM was 32.2% (95% CI, 18.5%-44.2%), indicating that approximately one third of deaths were statistically attributable to the observed distribution of CKM stages under the assumptions of the PAF model.

The stage allocation of years of life lost was sensitive to age but not to sex. Using age-specific shares alone gave 4.1%, 9.4%, 34.4%, 21.6%, and 30.6% of years of life lost across stages 0 to 4, against 4.0%, 9.4%, 34.6%, 21.6%, and 30.4% when both age and sex were used. Ignoring age entirely shifted 5.5 percentage points of the total onto stage 4, confirming that age is the dominant driver of the stage distribution of death (Table S6).

YLD rates differed by race and ethnicity (Table S7): 2,433 per 100,000 among non-Hispanic Black adults, 35% above non-Hispanic White adults (1,796), with intermediate rates in Hispanic adults (1,443) and the lowest rates among Asian and other adults (1,307). Stage 2 accounted for 55.1% of non-Hispanic Black adults, compared with 47.2% of White adults.

Most adults in stage 3 entered through the risk criterion rather than the kidney criterion: 80.1% met the PREVENT threshold without high-risk kidney disease, 10.7% met both, and 9.2% entered through high-risk kidney disease alone. The kidney-only group was much younger (mean age 52.7 versus 75.5 years), so this small subset identifies adults more than two decades earlier in the life course than the risk-based majority.

Age-standardized to the 2020 US population, the overall DALY rate was 7,881 per 100,000 (crude 8,537), and the stage distribution was 13.9%, 21.3%, 48.3%, 5.5%, and 8.4% for stages 0 to 4, against crude values of 13.8%, 21.3%, 49.1%, 6.5%, and 9.3%. Standardization therefore attenuated the apparent burden of the advanced stages.

Burden was higher in men than in women (10,291 versus 6,851 DALYs per 100,000), and men accounted for 59.1% of total DALYs despite a slightly smaller population. DALYs per 100,000 rose from 938 at 20-24 years to 27,013 at 75-84 years (Table 5).

### Adjusted mortality gradient

Unadjusted mortality rose steeply across stages (287, 390, 1,123, 6,709, and 6,375 per 100,000 person-years for stages 0-4), but most of that gradient was attributable to age. After adjustment, HRs relative to stage 0 were 0.99 (95% CI, 0.78-1.25) for stage 1, 1.39 (1.13-1.71) for stage 2, 2.21 (1.78-2.75) for stage 3, and 3.22 (2.60-3.99) for stage 4 (Table 3, Figure 3). Stages 2 through 4 were each significantly elevated; stage 1 was not.

**Figure 3.**
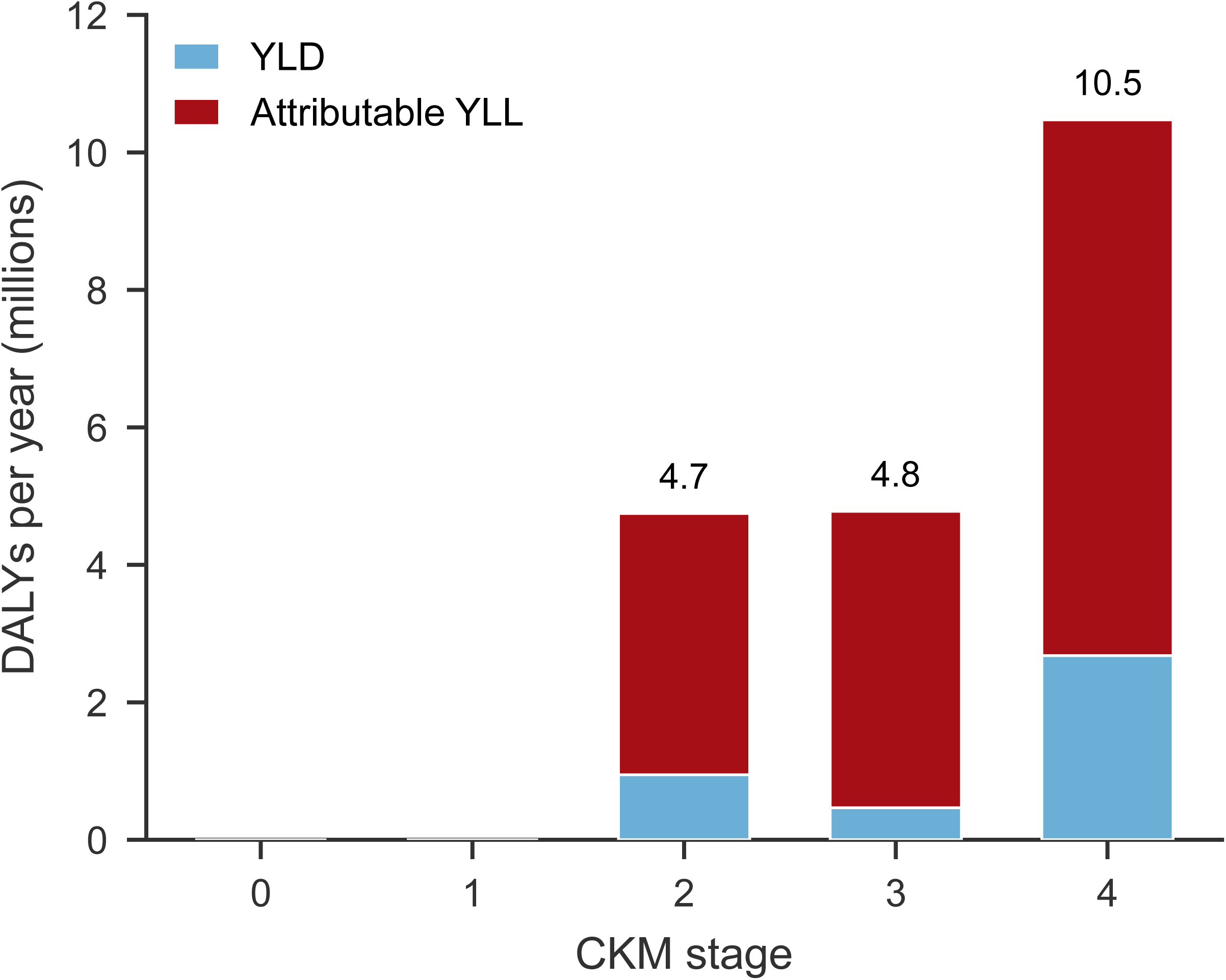
Adjusted hazard ratios for all-cause mortality by CKM stage, relative to stage 0. The model was adjusted for age, sex, and race and fitted to 27,012 participants and 4,706 deaths in the pooled 1999-2018 linked mortality cohort. Error bars indicate 95% confidence intervals; the x axis is logarithmic.

**Table 3.** Adjusted hazard ratios for all-cause mortality and population attributable fraction.

| **Stage** | **Deaths, n** | **Person-years** | **Crude rate per 100,000** | **Adjusted HR (95% CI)** | **Attributable YLL, %** |
|---|---|---|---|---|---|
| 0 | 99 | 34,483 | 287 | 1.00 (reference) | 0.0 |
| 1 | 233 | 59,763 | 390 | 0.99 (0.78-1.25) | 0.0 |
| 2 | 1,484 | 132,123 | 1,123 | 1.39 (1.13-1.71) | 23.9 |
| 3 | 1,333 | 19,867 | 6,709 | 2.21 (1.78-2.75) | 27.1 |
| 4 | 1,557 | 24,427 | 6,375 | 3.22 (2.60-3.99) | 49.0 |

Although crude mortality rates were similar in stages 3 and 4 (6,709 versus 6,375 per 100,000 person-years), the adjusted hazard ratio was lower in stage 3 than in stage 4 (2.21 versus 3.22). This finding suggests that the PREVENT-based criterion identifies a population with a high absolute short-term mortality burden, while its age-, sex-, and race-adjusted relative hazard remains lower than that of adults with established cardiovascular disease.

### Cause-specific cardiovascular mortality

The pattern held for cardiovascular death specifically. Crude cardiovascular mortality was 35 per 100,000 person-years in stage 0, 72 in stage 1, 294 in stage 2, 2,230 in stage 3, and 2,632 in stage 4, so absolute cardiovascular mortality in stage 3 approached that in stage 4 while both were more than sixty-fold higher than in stage 0. After adjustment for age, sex, and race, cause-specific hazard ratios were 1.41 (95% CI, 0.74-2.68) for stage 1, 2.63 (1.48-4.69) for stage 2, 4.73 (2.62-8.54) for stage 3, and 9.00 (5.03-16.13) for stage 4 (Table S8). This pattern reflects the older age structure of stage 3: in absolute terms a stage 3 adult faced a cardiovascular death rate about 85% that of a stage 4 adult, while the adjusted relative hazard remained lower.

### Treatment intensity despite similar crude mortality

Adults in stage 3 had mortality close to that of stage 4 but received less intensive preventive treatment (Table 6). Blood pressure was at or above 130/80 mm Hg in 65.6% of adults in stage 3 and 50.1% in stage 4; low-density lipoprotein cholesterol was at or above 70 mg/dL in 78.4% and 64.5%, respectively. Lipid-lowering therapy was absent in 53.7% of adults in stage 3 compared with 33.6% in stage 4, a difference of 20.1 percentage points, whereas antihypertensive use was similar in the two stages (65.4% versus 62.5%). The gap was therefore concentrated in lipid management, not in blood pressure treatment.

Treatment intensity improved in both stages over the study period, but the gap between them did not close (Figure 5). The proportion of adults in stage 3 taking no lipid-lowering drug fell from 73.4% in 2005-2006 to 53.7% in 2021-2023, and the corresponding proportion in stage 4 fell from 54.1% to 33.6%. The absolute difference was 19.3 percentage points at the start of the period and 20.1 percentage points at the end. The proportion with low-density lipoprotein cholesterol at or above 70 mg/dL moved in parallel, from 92.7% to 78.4% in stage 3 and from 79.6% to 64.5% in stage 4, a gap that remained between 13 and 14 percentage points throughout.

### Two-decade trend

Between 2006 and 2016, with a constant blood pressure protocol, stage 0 prevalence declined by 2.28% per year (95% CI, -3.36 to -1.19) and stage 1 rose by 3.11% per year (1.89 to 4.35); stage 2 declined modestly (-0.85% per year) and stages 3 and 4 were unchanged (0.06% and 0.27% per year, both non-significant) (Figure 1; Table S3). Joinpoint regression placed breakpoints at 2016 for stages 0, 1, and the composite of stages 2 and above, coinciding with the change in blood pressure instrumentation; the apparent reversal after 2016 should not be interpreted epidemiologically.

Because the stage-specific mortality gradient did not change across the study period (likelihood ratio test for stage-by-period interaction, P=0.63), stage-specific HRs estimated from 2005-2018 were applied to 2021-2023 death counts.

### Burden attributable to modifiable exposures

In the 16,220 participants with complete exposure and covariate data (1,767 deaths), current smoking carried an adjusted hazard ratio of 1.90 (95% CI, 1.68-2.15) and physical inactivity 1.65 (1.49-1.84) for all-cause mortality (Table 4). In 2017-March 2020, 16.2% of adults smoked, 44.8% were physically inactive, and 9.0% had both exposures.

**Table 4.** Adjusted hazard ratios and population attributable fractions for modifiable exposures outside the CKM definition.

| **Exposure** | **Prevalence, %** | **Adjusted HR (95% CI)** | **PAF, %** |
|---|---|---|---|
| Current smoking | 16.2 | 1.90 (1.68-2.15) | 12.7 |
| Physical inactivity | 44.8 | 1.65 (1.49-1.84) | 22.6 |
| Joint | 52.0 | Not estimated | 30.5 (25.7-35.1) |

*Hazard ratios adjusted for age, sex, race, family income-to-poverty ratio, education, and body mass index.*
*Prevalences are from NHANES 2017-March 2020. The joint distribution was 48.1% with neither exposure, 7.2% with smoking only, 35.8% with physical inactivity only, and 9.0% with both. PAF indicates population attributable fraction.*

**Table 5.** DALYs by sex and age group, 2021-2023.

| Group | YLD | Attributable YLL | DALYs | DALYs per 100,000 | % of total |
|---|---|---|---|---|---|
| Men | 2,387,569 | 9,428,647 | 11,816,216 | 10,291 | 59.1 |
| Women | 1,705,725 | 6,456,353 | 8,162,078 | 6,851 | 40.9 |
| 20-24 years | 40,101 | 127,425 | 167,526 | 938 | 0.8 |
| 25-34 years | 131,095 | 612,531 | 743,626 | 1,661 | 3.7 |
| 35-44 years | 179,698 | 896,703 | 1,076,401 | 2,601 | 5.4 |
| 45-54 years | 495,290 | 1,728,345 | 2,223,634 | 6,369 | 11.1 |
| 55-64 years | 1,067,694 | 3,524,276 | 4,591,971 | 10,676 | 23.0 |
| 65-74 years | 1,116,346 | 4,683,370 | 5,799,716 | 18,052 | 29.0 |
| 75-84 years | 1,063,071 | 4,312,350 | 5,375,421 | 27,013 | 26.9 |

*Percentages for the age rows sum to 100% across ages 20-84; NHANES top-codes age at 80 years, so no participants contribute to an 85 years and older stratum in 2021-2023.*

**Table 6.** Treatment intensity in CKM stages 2 to 4, 2021-2023.

| **Measure** | **Stage 2** | **Stage 3** | **Stage 4** |
|---|---|---|---|
| Adults, millions | 114.8 | 15.3 | 21.7 |
| Blood pressure >=130/80 mm Hg, % | 55.1 | 65.6 | 50.1 |
| No antihypertensive medication, % | 70.9 | 34.6 | 37.5 |
| LDL cholesterol >=70 mg/dL, % | 90.5 | 78.4 | 64.5 |
| No lipid-lowering medication, % | 76.4 | 53.7 | 33.6 |

*Values are survey-weighted percentages of adults in each stage. Stage 3 comprises adults without clinical cardiovascular disease who meet the PREVENT risk criterion or have high-risk chronic kidney disease; stage 4 comprises adults with self-reported cardiovascular disease.*

Current smoking accounted for 12.7% of CKM DALYs and physical inactivity for 22.6%. Because the two exposures co-occur, their joint attributable fraction was 30.5% (95% CI, 25.7-35.1), corresponding to 6.1 million DALYs per year (Figure 4). Physical inactivity therefore contributed roughly twice as much burden as smoking.

**Figure 4.**
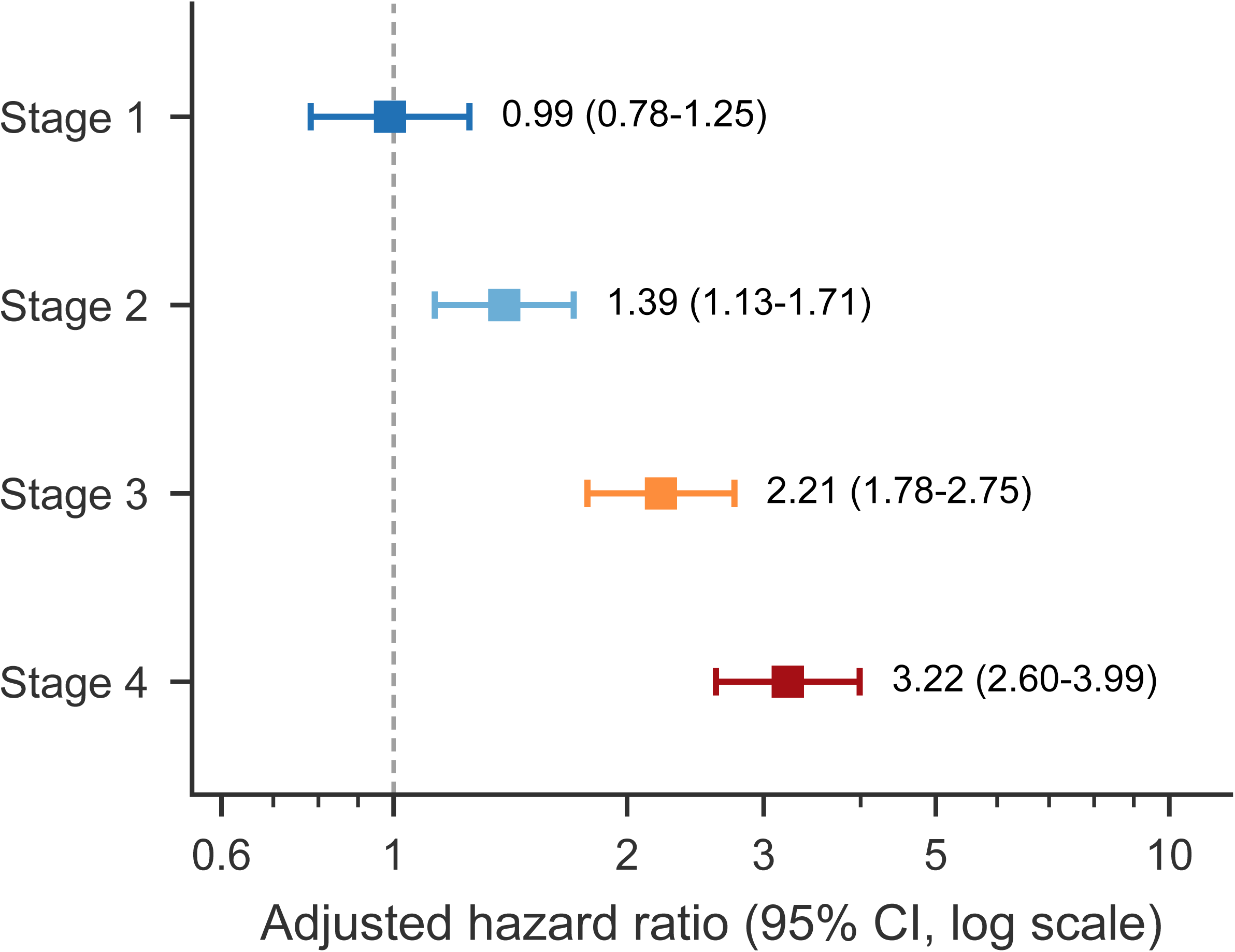
Population attributable fractions for current smoking and physical inactivity, estimated in the 2005-2018 linked mortality cohort and applied to 2017-March 2020 exposure prevalences. The joint estimate allows for the co-occurrence of the two exposures and is therefore smaller than the sum of the individual fractions. Error bar indicates the 95% confidence interval for the joint estimate.

**Figure 5.**
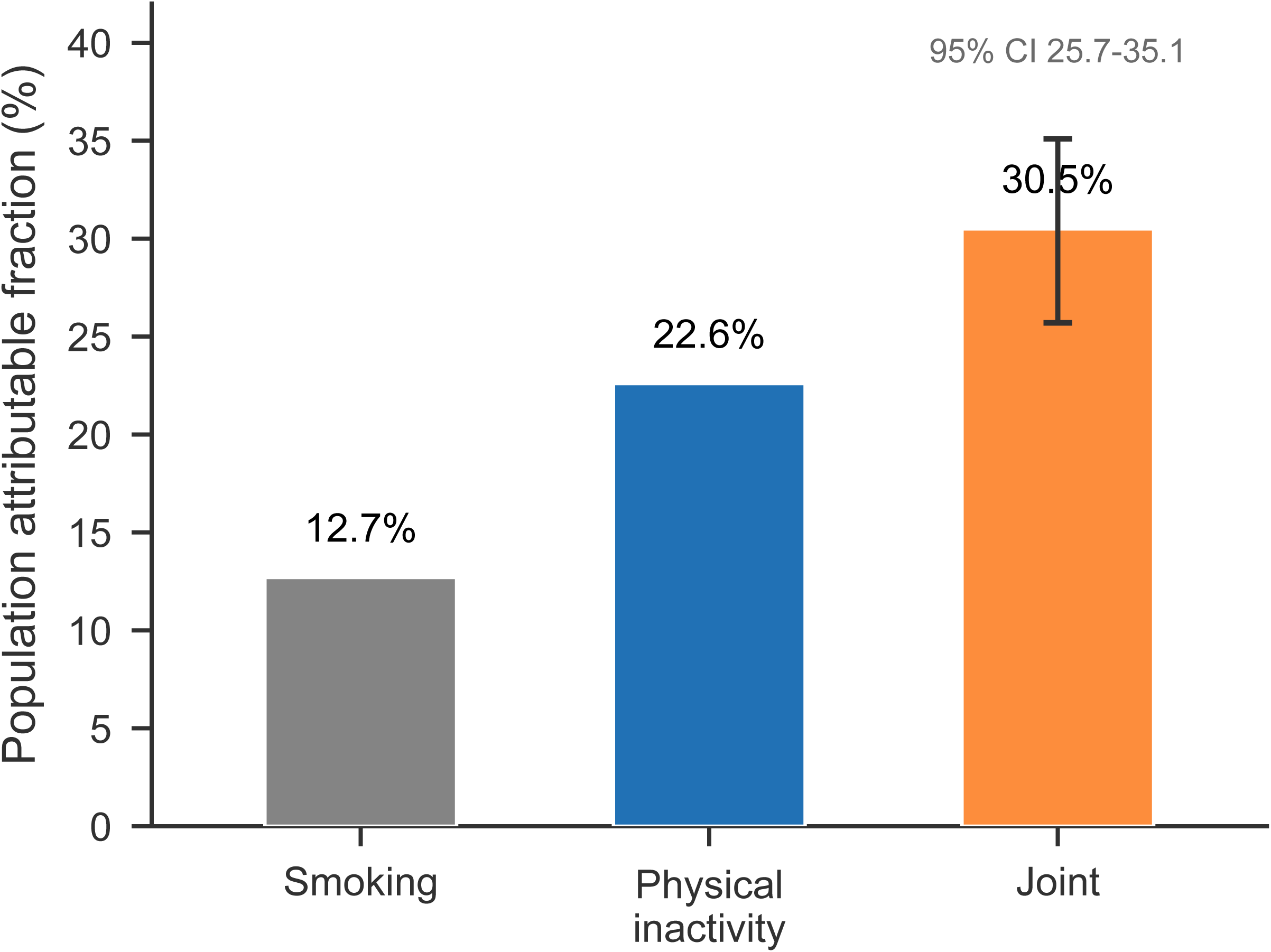
Persistence of the lipid-lowering gap between CKM stages 3 and 4, 2005 to 2023. A, Proportion of adults taking no lipid-lowering medication. B, Proportion with low-density lipoprotein cholesterol at or above 70 mg/dL. Both stages improved over time at similar rates, so the absolute difference between them was unchanged. The 2017-2018 cycle is omitted because it overlaps the 2017-March 2020 cycle.

### Sensitivity analyses

Raising the PREVENT threshold to 30% reduced stage 3 prevalence from 6.5% to 2.1% but changed the PAF by only about 2 percentage points. Omitting the low-density lipoprotein criterion moved 2.8 percentage points from stage 2 to stages 0-1 and changed the PAF to 33.0%. Relaxing the stage 0 completeness rule had no effect, because all stage 0 participants in 2021-2023 already had complete core inputs. Assigning all conditions the mild or severe end of the GBD severity range moved YLD from 4.09 million to 2.97 million and 9.95 million, respectively, corresponding to DALYs of 19.5 to 26.5 million per year. The Monte Carlo 95% confidence interval for total DALYs was 8.9 to 29.8 million per year, dominated by uncertainty in the stage-specific hazard ratios, not in prevalence (Table S4).

## Discussion

In this nationally representative analysis, a staging framework built on predicted risk identified a population whose crude mortality was close to that of established cardiovascular disease, and CKM syndrome as a whole accounted for approximately 20.0 million DALYs per year among US adults, or 7,881 age-standardized DALYs per 100,000. The burden is concentrated in stages 3 and 4, which comprise 15.8% of adults but generated 76% of attributable years of life lost. Over the same two decades, however, stage 1 expanded by 3.1% per year and stage 0 contracted by 2.3% per year, with no change in stages 3 and 4.

This pattern has implications for how prevention is evaluated. Stage 1 was not associated with a statistically significant excess in adjusted mortality (HR, 0.99; 95% CI, 0.78-1.25), whereas stage 2 was associated with a moderate increase (HR, 1.39; 95% CI, 1.13-1.71). A prevention program evaluated solely by short-term deaths averted may show little immediate mortality benefit among the 21% of adults classified as stage 1. However, the absence of a statistically significant excess in short-term mortality does not imply an absence of preventive benefit. The principal value of intervention at this stage may be preventing progression to stages 3 or 4, an outcome that this cross-sectional analysis cannot observe. Burden metrics therefore tend to favor late-stage treatment, potentially more so for CKM than for conditions whose burden is concentrated earlier.

Crude mortality in stage 3 was close to that in stage 4, although the adjusted hazard ratio remained lower. Stage 3 is defined by predicted risk rather than observed disease, and its members were older and more likely to have chronic kidney disease. The finding that a risk equation identifies a group with crude short-term mortality similar to that of adults with established cardiovascular disease supports the construct validity of the staging framework and suggests that risk-based staging adds information beyond diagnosis counts.

Our estimates can be compared with existing CKM research. Hyun and colleagues reported stage distribution and risk-factor control gaps for 2021-2023 but did not estimate health loss.2 Our stage prevalences (13.8%, 21.3%, 49.1%, 6.5%, and 9.3%) differed from theirs by no more than 3.0 percentage points, despite differences in the operational definitions of hypertension and the PREVENT threshold. This concordance supports the robustness of the underlying construct. Their analysis was confined to a single cycle and did not address mortality, time trends, or attributable burden. They also documented treatment gaps across stages 2 to 4; our treatment analysis differs by anchoring the gap to the mortality comparison between stages 3 and 4. Kibbi and colleagues examined albuminuria within CKM stages,6 and Zuin and Piazza examined pulmonary embolism mortality in CKM,7 but neither quantified overall burden.

The 80% share of YLL in total DALYs contrasts with many chronic conditions in which YLD dominates. This reflects the lethality of the constituent diseases rather than measurement artifact: cardiovascular disease and chronic kidney disease are leading causes of death, and their disability weights, while non-trivial, are modest relative to the years of life lost.

The analysis has several strengths. Risk factors were measured rather than self-reported, staging incorporated predicted risk rather than diagnosis alone, and disability weights were drawn from GBD health states with severity and anemia stratification rather than applied uniformly. The mortality model included 27,012 participants and 4,706 deaths followed for up to two decades, allowing the stage gradient to be estimated with reasonable precision and its stability over time to be assessed.

Direct comparison with GBD estimates is not possible because GBD does not report burden by CKM stage. The magnitude is nevertheless plausible: the syndrome’s constituent conditions, including ischemic heart disease, stroke, diabetes, and chronic kidney disease, are among the leading contributors to DALYs in the United States. Our estimates aggregate these conditions through a single staged framework instead of treating them as separate cause categories. A recent GBD-based comparison of CKM burden across seven countries used DALY rates for ischemic heart disease, diabetic kidney disease, and fatty liver disease rather than stage-specific estimates and therefore cannot serve as a numerical benchmark; no stage-specific DALY estimate exists for the United States.16

Two exposures that sit outside the CKM definition, current smoking and physical inactivity, accounted for almost a third of the syndrome’s burden. CKM staging describes who is at risk but not what is driving the risk. Smoking and inactivity are causes, not stages, and they can be modified at the population level without the clinical infrastructure required for stage 3 and 4 care.

The difference between the two exposures is relevant to prevention policy. Physical inactivity contributed roughly twice the burden of smoking (22.6% versus 12.7%), despite tobacco control having a much more developed policy architecture. Physical activity has been less institutionalized within cardiovascular prevention, and these estimates suggest that its share of the CKM agenda should reflect its contribution.

For clinicians, the implication is narrower than the policy one. Adults who meet the PREVENT threshold for stage 3 had crude short-term mortality similar to that of adults with established cardiovascular disease, yet the data suggest that they are treated more like a high-risk primary-prevention population than a secondary-prevention population. The treatment gap we observed was concentrated in lipid lowering: approximately half of adults in stage 3 were taking no lipid-lowering medication, and four in five had low-density lipoprotein cholesterol at or above 70 mg/dL, compared with one in three and two in three, respectively, in stage 4. Closing this gap does not require a new risk tool, because the group is already identified by the existing one.

Our results should be interpreted in light of several limitations. First, NHANES is cross-sectional; stage at a single examination is used as a proxy for a lifelong trajectory, and progression cannot be observed. Second, the PAF is imprecise (95% CI, 18.5%-44.2%) because the linked mortality cohort contains 4,706 deaths across five stages; the descriptive stage-specific burden estimates do not share this limitation. Third, stage-specific HRs from 2005-2018 were applied to 2021-2023 death counts. Although the absence of a stage-by-period interaction supports this approach, secular changes in treatment could still have altered absolute risks. Fourth, the transition from mercury to oscillometric blood pressure measurement in 2017 creates a discontinuity that precludes trend inference after 2016. Fifth, disability weights for ischemic heart disease and stroke were based on GBD 2013 severity distributions because updated distributions were not publicly accessible; the mild and severe bounds we report span the plausible range. Sixth, the stage-based PAF is a model-based attribution, not the causal effect of an intervention that would move every adult to stage 0, and it assumes that the stage-specific hazard ratios are unconfounded. Seventh, the DALY estimates assume a steady state; a cross-sectional design cannot capture the lag between exposure and death, so the burden associated with today’s stage 1 will become apparent only in future estimates.

These limitations do not affect the central inference, which rests on the shape of the burden distribution rather than its absolute level; every sensitivity analysis preserved the concentration of burden in stages 3 and 4.

## Conclusions

CKM syndrome accounts for approximately 20.0 million DALYs annually among US adults, with the burden concentrated in the 15.8% of adults at stages 3 and 4. Excess mortality begins at stage 2 and increases through stages 3 and 4. Yet the population has been moving into stage 1 for two decades, where adjusted mortality was not significantly higher than in adults with no risk factors. Closing the gap between where burden is measured and where it is being generated is the central challenge the CKM framework now faces.

## Data Availability

The data analysed in this study are publicly available and were not generated by the authors. NHANES data were obtained from the National Center for Health Statistics (https://www.cdc.gov/nchs/nhanes/). Public-use linked mortality files were obtained from the NCHS Data Linkage Program (https://ftp.cdc.gov/pub/Health_Statistics/NCHS/datalinkage/linked_mortality/). National death counts were obtained from CDC WONDER (https://wonder.cdc.gov/), and period life tables from the National Center for Health Statistics (https://www.cdc.gov/nchs/products/life_tables.htm). Analysis code is available from the corresponding authors on reasonable request.

https://www.cdc.gov/nchs/nhanes/

## Acknowledgments

The authors thank the participants and staff of the National Health and Nutrition Examination Survey and the National Center for Health Statistics for making the data publicly available.

Use of Artificial Intelligence: ChatGPT (OpenAI) was used solely to improve the clarity and readability of the English text. It was not used to generate data, perform analyses, or interpret results. All authors reviewed and edited all content and take full responsibility for the manuscript.

## Data Availability Statement

NHANES data are publicly available from the National Center for Health Statistics. Linked mortality files are available from https://ftp.cdc.gov/pub/Health_Statistics/NCHS/datalinkage/linked_mortality/. Analysis code is available from the corresponding author on reasonable request.

## Nonstandard Abbreviations and Acronyms

CKM: cardiovascular-kidney-metabolic
DALY: disability-adjusted life year
YLD: years lived with disability
YLL: years of life lost
PAF: population attributable fraction
eGFR: estimated glomerular filtration rate
UACR: urine albumin-to-creatinine ratio
LMF: linked mortality file
NHANES: National Health and Nutrition Examination Survey
PREVENT: Predicting Risk of Cardiovascular Disease Events.

## Supplemental Material

Figure S1. Study design and analytic samples. Table S1. Operational definitions of CKM stages. Table S2. Cross-cycle variable harmonization. Table S3. Stage distribution by cycle. Table S4. Sensitivity analyses. Table S5. Disability weights applied. Table S6. Sensitivity of the stage allocation of years of life lost. Table S7. Stage distribution and years lived with disability by race and ethnicity. Table S8. Cause-specific cardiovascular mortality by CKM stage.

---

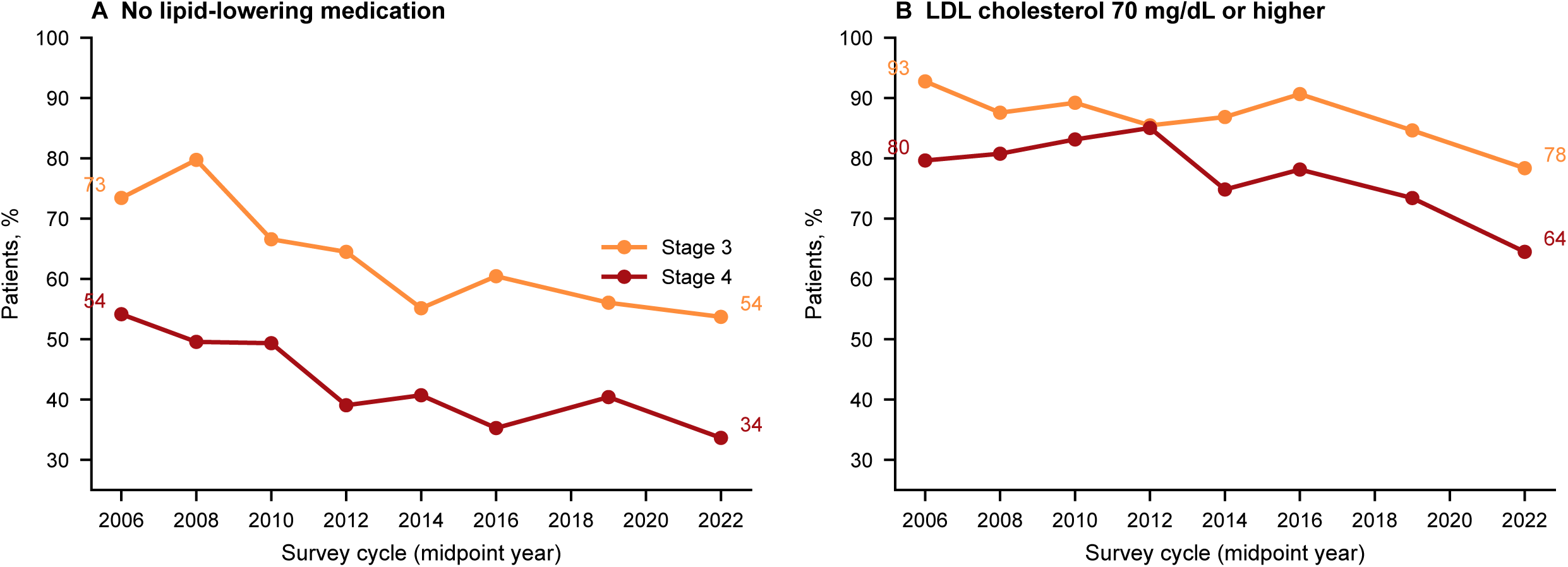
Central Illustration. Where CKM burden sits and how treatment differs across stages. A, Distribution of US adults across CKM stages 0 to 4, 2021-2023. B, Share of adults (upper bar) and share of attributable years of life lost (lower bar) by stage; stages 3 and 4 comprise 15.8% of adults but generate 76% of attributable years of life lost. C, Treatment intensity in stages 3 and 4, showing a lipid-lowering gap despite comparable mortality; the gap was 19.3 percentage points in 2005-2006 and 20.1 percentage points in 2021-2023. D, Composition of stage 3, with mean age shown above each bar: 80.1% entered through the PREVENT criterion alone, 10.7% through both criteria, and 9.2% through high-risk kidney disease alone. Colors denote stages 0 to 4 in panels A to C.

