## Supplemental Tables S1-S8 for "Risk-Based Cardiovascular-Kidney-Metabolic Staging Identifies a Population With Mortality Comparable to Established Cardiovascular Disease: Disability-Adjusted Life Years Across AHA Stages, 2005 to 2023"

*The supplementary material includes Figure S1 and Tables S1 to S8. Figure S1 is also submitted as a separate file.*

#### Table S1. Operational definitions of CKM stages

| Stage | Definition in this analysis |
|---|---|
| 0 | No CKM risk factor, and all five core inputs observed (body mass index, blood pressure, hemoglobin A1c, eGFR, UACR) |
| 1 | Overweight (BMI 25.0-29.9), obesity (BMI >=30), abdominal obesity (waist >=102 cm men, >=88 cm women), or prediabetes (HbA1c 5.7%-6.4% without diabetes) |
| 2 | Hypertension, diabetes, chronic kidney disease, hypertriglyceridemia (TG >=150 mg/dL), metabolic syndrome (NCEP ATP III), or LDL cholesterol >=160 mg/dL |
| 3 | No clinical CVD, and 10-year PREVENT total cardiovascular risk >=20% or high-risk CKD (eGFR <45 mL/min/1.73 m2 or UACR >=300 mg/g) |
| 4 | Self-reported congestive heart failure, coronary heart disease, angina, myocardial infarction, or stroke |

*Hypertension: measured systolic >=130 or diastolic >=80 mm Hg, or current antihypertensive medication; self-report of a prior diagnosis was not used. Diabetes: self-report, glucose-lowering medication, or HbA1c >=6.5%. CKD: eGFR <60 (2021 CKD-EPI) or UACR >=30 mg/g. Stages were assigned hierarchically, highest stage first.*

#### Table S2. Cross-cycle variable harmonization and file inventory

| Component | 1999-2000 | 2001-2002 | 2003-2004 | 2005-2018 | 2017-Mar 2020 | 2021-2023 |
|---|---|---|---|---|---|---|
| Demographics | DEMO | DEMO_B | DEMO_C | DEMO_[D-J] | P_DEMO | DEMO_L |
| Biochemistry | LAB18 | L40_B (no creatinine) | L40_C | BIOPRO | P_BIOPRO | BIOPRO_L |
| HbA1c | LAB10 | L10_B | L10_C | GHB | P_GHB | GHB_L |
| Total cholesterol, HDL | LAB13 (LBDHDL) | L13_B (LBDHDL) | L13_C (LBXHDD) | TCHOL + HDL (LBDHDD) | P_TCHOL + P_HDL | TCHOL_L + HDL_L (LBXHDD) |
| Triglycerides, LDL | LAB13AM (LBXTR) | L13AM_B | L13AM_C | TRIGLY (LBXTR) | P_TRIGLY | TRIGLY_L (LBXTLG) |
| Urine albumin, creatinine | LAB16 | L16_B | L16_C | ALB_CR | P_ALB_CR | ALB_CR_L |
| Hemoglobin | LAB25 | L25_B | L25_C | CBC | P_CBC | CBC_L |
| Blood pressure | BPX (mercury) | BPX_B | BPX_C | BPX (mercury) | BPXO (oscillometric) | BPXO_L (oscillometric) |
| Physical activity | PAQ (PAD200/PAD320) | PAQ_B | PAQ_C | PAD200/320 to 2006; PAQ650/665 from 2007 | P_PAQ (PAQ650/665) | PAQ_L (PAD790 series; not harmonised) |
| Questionnaires | BPQ, DIQ, MCQ, SMQ | _B suffix | _C suffix | _D to _J suffix | P_ prefix | _L suffix |
| Examination weight | WTMEC4YR | WTMEC4YR | WTMEC2YR | WTMEC2YR | WTMECPRP | WTMEC2YR |
| Fasting weight | WTSAF4YR | WTSAF4YR | WTSAF2YR | WTSAF2YR | WTSAFPRP | WTSAF2YR |

Legend. Cell entries are file stems; the variable suffix follows the cycle. Lipid variable names varied across cycles: LBDHDL (1999-2002), LBXHDD (2003-2004), LBDHDD (2005-2018), and LBXHDD again from 2021. Triglycerides are LBXTR through 2018 and LBXTLG from 2021.

*The 1999-2000 and 2003-2004 cycles were not combined with the intervening 2001-2002 cycle, and no cycle-spanning weight was constructed. They contributed to the unweighted Cox model only: serum creatinine is available in both cycles (LAB18 and L40_C), so CKM stage could be assigned, and no survey weight was applied in that model. The 2001-2002 cycle was excluded from all analyses because serum creatinine is absent from its public biochemistry file (L40_B), which precludes eGFR and therefore CKM staging. All weighted estimates (stage prevalence, YLD) were restricted to 2005-2006 onward, where the fasting subsample weights are available. 1999-2000 and 2003-2004 contributed to the mortality model only; stage prevalence and YLD were estimated from 2005-2023, where triglycerides are available for the full fasting subsample.*

*Variables extracted from the legacy files were: RIDAGEYR, RIAGENDR, RIDRETH1, RIDSTATR, DMDEDUC2, INDFMPIR (demographics); BMXBMI, BMXWAIST; BPXSY1-4, BPXDI1-4 (1999-2004) or BPXOSY1-3, BPXODI1-3 (2017 onwards); LBXSCR,*

*LBXSGL; LBXGH; LBXTC, LBDHDL/LBXHDD; LBXTR, LBDLDL; URXUMA, URXUCR; LBXHGB; BPQ020, BPQ050A, BPQ100D; DIQ010, DIQ050, DIQ070 (DID070 in 2005-2008); MCQ160B-F; SMQ020, SMQ040; PAD200, PAD320, PAQ650, PAQ665.*

### Table S3. Stage distribution by survey cycle

| Cycle | n | Stage 0 | Stage 1 | Stage 2 | Stage 3 | Stage 4 | >= Stage 2 |
|---|---|---|---|---|---|---|---|
| 2005-06 | 2130 | 15.1 | 17.7 | 52.0 | 6.0 | 9.2 | 67.1 |
| 2007-08 | 2589 | 15.2 | 20.3 | 49.5 | 6.1 | 9.0 | 64.5 |
| 2009-10 | 2742 | 14.2 | 22.8 | 48.8 | 6.0 | 8.2 | 63.1 |
| 2011-12 | 2417 | 12.9 | 22.1 | 50.2 | 5.2 | 9.6 | 65.0 |
| 2013-14 | 2537 | 13.7 | 23.0 | 48.4 | 6.1 | 8.9 | 63.3 |
| 2015-16 | 2234 | 12.0 | 26.1 | 46.6 | 6.3 | 8.9 | 61.9 |
| 2017-18 | 2371 | 12.6 | 21.8 | 49.1 | 6.4 | 10.1 | 65.5 |
| 2017-Mar20 | 3874 | 12.2 | 23.2 | 48.6 | 5.7 | 10.3 | 64.6 |
| 2021-23 | 3057 | 13.8 | 21.3 | 49.1 | 6.5 | 9.3 | 64.9 |

*Values are survey-weighted percentages. 2017-2018 is shown for internal consistency but excluded from trend analyses because it overlaps 2017-March 2020.*

### Table S4. Sensitivity analyses

| Analysis | Result |
|---|---|
| Base case | PAF 32.2% (95% CI 18.5-44.2); 20.0 million DALYs per year |
| PREVENT threshold 30% instead of 20% | Stage 3 prevalence 6.5% to 2.1%; PAF changed by about 2 percentage points |
| LDL >=160 mg/dL omitted from stage 2 | 2.8 percentage points moved from stage 2 to stages 0-1; PAF 33.0% |
| Stage 0 completeness rule relaxed | No change (all stage 0 participants had complete core inputs in 2021-2023) |
| All disability weights at the mild end | YLD 2.97 million; DALYs 19.5 million per year |
| All disability weights at the severe end | YLD 9.95 million; DALYs 26.5 million per year |
| Monte Carlo (4000 draws) | PAF 9.8-52.1 in the 7-cycle cohort; 18.5-44.2 with 1999-2004 added |

### Table S5. Disability weights applied

| Condition | GBD health state | Weight |
|---|---|---|
| Diabetes | Uncomplicated diabetes mellitus | 0.049 |
| Ischemic heart disease | Severity-weighted (GBD 2013 Table 2) | 0.0731 |
| Stroke, chronic | Severity-weighted (GBD 2013 Table 2) | 0.0845 |
| Heart failure | Severity-weighted (GBD 2023 Table S3) | 0.0923 |
| CKD stage 3, no anemia | Asymptomatic | 0.000 |
| CKD stage 3, mild / moderate / severe anemia | GBD CKD Table 7 | 0.004 / 0.052 / 0.149 |
| CKD stage 4, no anemia | GBD CKD Table 7 | 0.104 |
| CKD stage 4, mild / moderate / severe anemia | GBD CKD Table 7 | 0.108 / 0.150 / 0.237 |
| CKD stage 5 (dialysis) | End-stage renal disease on dialysis | 0.569 |

*Comorbid conditions were combined multiplicatively: DW = 1 - product(1 - DWi). Among participants with chronic kidney disease in 2021-2023, 75.5% had no anemia, 17.4% mild anemia, 6.7% moderate anemia, and 0.4% severe anemia; hemoglobin was missing for 0.5% of the analytic sample.*

### Table S6. Sensitivity of the stage allocation of years of life lost

| Source of stage-specific death shares | Stage 0 | Stage 1 | Stage 2 | Stage 3 | Stage 4 | Cumulative YLL 2021-2023, millions |
|---|---|---|---|---|---|---|
| Age and sex (base case) | 4.0 | 9.4 | 34.6 | 21.6 | 30.4 | 148.2 |
| Age only | 4.1 | 9.4 | 34.4 | 21.6 | 30.6 | 148.2 |
| Sex only | 2.2 | 5.5 | 27.4 | 29.2 | 35.8 | 148.2 |
| Neither (pooled) | 2.2 | 5.5 | 27.3 | 29.2 | 35.9 | 148.2 |

*Values are the percentage of total years of life lost allocated to each stage. The total is invariant because the shares sum to one within each stratum; the choice of shares therefore affects the distribution of burden across stages, not its magnitude. Ignoring age moves 5.5 percentage points of burden into stage 4, whereas adding sex to age changes the allocation by less than 0.2 percentage points. A single-cycle (2017-2018) variant could not be estimated because several stage-by-age cells contained no deaths.*

#### Table S7. Stage distribution and years lived with disability by race and ethnicity, 2021-2023

| Group | Adults, millions | Stage 0 | Stage 1 | Stage 2 | Stage 3 | Stage 4 | YLD | YLD per 100,000 |
|---|---|---|---|---|---|---|---|---|
| Non-Hispanic White | 142.2 | 13.3 | 21.4 | 47.2 | 7.3 | 10.8 | 2,553,207 | 1,796 |
| Non-Hispanic Black | 25.5 | 10.8 | 18.7 | 55.1 | 6.6 | 8.8 | 619,617 | 2,433 |
| Hispanic | 39.3 | 12.2 | 27.5 | 49.5 | 5.2 | 5.6 | 567,337 | 1,443 |
| Asian and other | 27.0 | 21.6 | 14.7 | 52.7 | 4.2 | 6.8 | 353,133 | 1,307 |

*Stage values are survey-weighted percentages of adults in each group and sum to 100% across stages. YLD indicates years lived with disability. Years of life lost and DALYs are not reported by race and ethnicity because the national mortality source used in this analysis does not provide death counts by race; YLD is derived entirely from survey data and is therefore not subject to that limitation.*

#### Table S8. Cause-specific cardiovascular mortality by CKM stage

| Stage | Participants | Cardiovascular deaths | Person-years | Deaths per 100,000 person-years | Adjusted HR (95% CI) |
|---|---|---|---|---|---|
| 0 | 3,003 | 12 | 34,483 | 35 | 1.00 (reference) |
| 1 | 5,410 | 43 | 59,763 | 72 | 1.41 (0.74-2.68) |
| 2 | 12,792 | 389 | 132,123 | 294 | 2.63 (1.48-4.69) |
| 3 | 2,584 | 443 | 19,867 | 2,230 | 4.73 (2.62-8.54) |
| 4 | 3,223 | 643 | 24,427 | 2,632 | 9.00 (5.03-16.13) |

Cardiovascular death was defined as an underlying cause of death of diseases of the heart or cerebrovascular disease in the public-use linked mortality files. Hazard ratios are from Cox models with age (per 10 years), sex, and race as covariates; deaths from other causes were censored. The analytic sample comprised 27,012 adults and 1,530 cardiovascular deaths from the 1999-2000, 2003-2004, and 2005-2018 cycles. This sample was identical to that used for the all-cause model, allowing direct comparison of the two sets of estimates.
