## Supplementary figures and images for "Risk-Based Cardiovascular-Kidney-Metabolic Staging Identifies a Population With Mortality Comparable to Established Cardiovascular Disease: Disability-Adjusted Life Years Across AHA Stages, 2005 to 2023"

### Figure S1

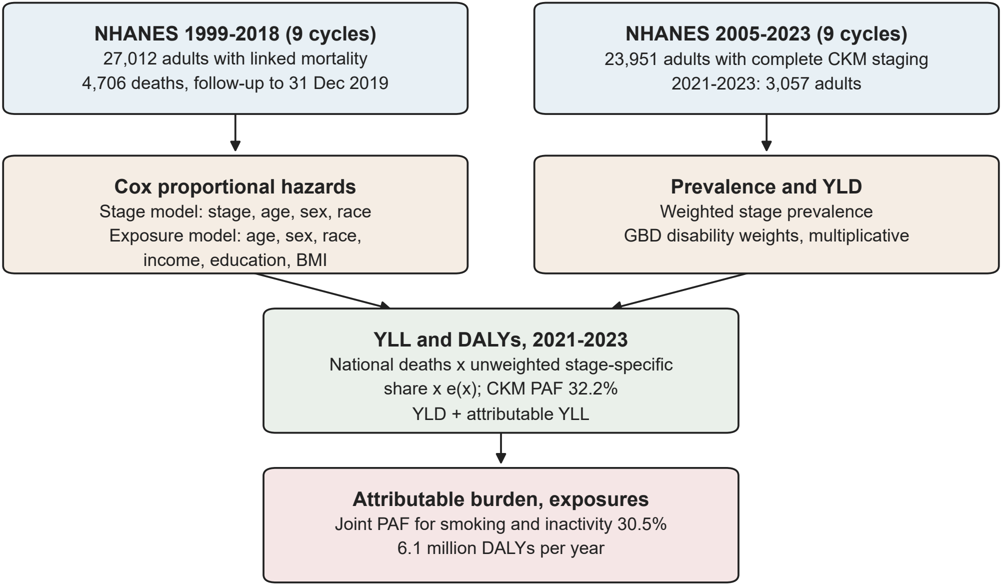


*CDC WONDER death counts (2021-2023) and NCHS 2024 period life tables*
